# Effectiveness of An Impedance Cardiography Guided Treatment Strategy to Improve Blood Pressure Control in A Real-World Setting: Results from A Pilot Pragmatic Clinical Trial

**DOI:** 10.1101/2021.04.09.21255233

**Authors:** Luyan Wang, Yuan Lu, Hongyi Wang, Jianlei Gu, Zheng J. Ma, Zheng Lian, Zhiying Zhang, Harlan M. Krumholz, Ningling Sun

## Abstract

**Background:** Hypertension is a hemodynamic-related disorder characterized by abnormalities of the cardiac output (CO) and/or systemic vascular resistance (SVR). We hypothesized that selecting antihypertensive therapy based on patients’ hemodynamic profile could improve blood pressure (BP) control more effectively than standard care in hypertensive patients in real-world clinical practice.

**Methods:** We conducted a pilot single-center, pragmatic randomized trial involving adults with uncontrolled hypertension who sought outpatient care at a hypertension clinic of the Peking University People’s Hospital, the largest teaching hospital of Peking University, in Beijing China, between December 2018 and December 2019. Participants were randomly assigned to the standard care group or the hemodynamic group in a 1:1 ratio. Impedance cardiography (ICG) was performed with all participants to measure hemodynamic parameters. Only physicians in the hemodynamic group were provided with patients’ ICG findings and a computerized clinical decision support of recommended treatment choices based on patients’ hemodynamic profiles. The primary outcomes were the changes in systolic BP (SBP) and diastolic BP (DBP) levels at the follow-up visit 4-12 weeks after baseline. Secondary outcomes included achievement of BP goal of <140/ 90 mmHg and the changes in BP by baseline BP, age, sex, and BMI.

**Results:** A total 102 adults (mean age was 54±14 years; 41% were women) completed the study. The mean baseline SBP was 150.9 (±11.5) mmHg and mean baseline DBP was 91.1 (±11.3) mmHg. At the follow-up visit, the mean SBP and DBP decreased by 19.9 and 11.3 mmHg in the hemodynamic group, as compared with 12.0 and 4.9 mmHg in the standard care group (P value for difference between groups <0.001 for both SBP and DBP). The proportion of patients achieving BP goal of <140/ 90 mmHg in the hemodynamic group was 67%, as compared with 41% in the standard care group (P=0.017). The hemodynamic group had a larger effect on BP reduction consistently across subgroups by age, sex, BMI, and baseline BP.

**Conclusions:** An ICG-guided treatment strategy led to greater reductions in BP levels than were observed with standard care in a real-world population of outpatients with hypertension. There is a need for further validation of this strategy for improving blood pressure treatment selection. (Funded by internal research grant from the Peking University People’s Hospital; ClinicalTrials.gov number: NCT04715698.)

## INTRODUCTION

Hypertension is a hemodynamic-related disorder characterized by abnormalities of the cardiac output (CO), systemic vascular resistance (SVR), or a combination of both.^1^ Despite that hypertension is routinely diagnosed and managed based on degree of blood pressure (BP) elevation alone, patients with similar degree of BP elevation can have different underlying hemodynamic profiles.^2, 3^ These variations in hemodynamic profiles may have important implications for treatment selection because the choice for patients with a higher CO might be different than for those with a higher SVR. Selecting treatment strategies based on hemodynamic profiles for patients with hypertension may improve BP control.

Impedance cardiography (ICG) is a safe and accurate non-invasive tool to measure hemodynamic parameters^4, 5^ that can be performed in the outpatient setting.^6, 7^ Measurement of the various hemodynamic components using ICG in stable patients with hypertension provides information that may enable more effective targeted drug management. Although several previous studies have used ICG to evaluate hemodynamic parameters and demonstrated that ICG-guided therapy improves BP control,^7-9^ they used a traditional randomized controlled trial design, in which the operationalization of the intervention had stricter instructions and patients were more frequently monitored than routine clinical care. Whether an ICG-guided strategy for hypertension treatment can lead to improvements in BP control in real-world clinical settings has been rarely tested. Additionally, previous studies were all conducted in the United States; no study has focused on low- and middle-income counties where healthcare resources are limited, patient characteristics and clinical practice patterns are different.

Accordingly, we conducted a pilot pragmatic randomized trial to produce preliminary data about the effectiveness of ICG-guided strategies for patients with hypertension in routine clinical care in China. We hypothesized that selecting antihypertensive therapy based on each patient’s hemodynamic profile measured by ICG could lead to more effective BP reduction and hypertension control than standard care in hypertensive patients in a real-world setting.

## METHODS

### Eligibility

The study population was patients who sought outpatient care for hypertension in the hypertension clinic of the Cardiology Department at the Peking University People’s Hospital between June and December 2019 in Beijing, China. Patients were eligible if they were 18 to 85 years old, were local residents, had a diagnosis of essential hypertension, and were currently on 0 to 3 antihypertensive medications with systolic blood pressure (SBP) of ≥140 mmHg or diastolic blood pressure (DBP) of ≥90 mmHg. Patients were excluded if they were already on more than 3 antihypertensive agents (considered as resistant hypertension); had on-site SBP of <140 mmHg and DBP of <90 mmHg; had secondary hypertension, severe renal disease, cancer, severe valvular disease, cerebrovascular event within 6 months, atrial fibrillation; or had uncontrolled diabetes with fasting blood glucose of 11.1 mmol/L. The study was reviewed and approved by the Peking University Institutional Review Board. All hypertensive outpatients provided written informed consent and had study procedures consistent with the protocol.

### Randomization and procedure

After informed consent, patients meeting inclusion/exclusion criteria were randomized in a 1:1 ratio to the hemodynamic group or the standard care group using a random number generator. Patients’ information including age, sex, weight, height, BP, and antihypertensive medications was collected by nurses during the outpatient visit. Weight was measured to the nearest 0.1 kg with patients wearing light indoor clothing and no shoes. Height was measured to the nearest 0.1 cm, using a portable stadiometer (Omron HNJ-318; Omron Corporation, Kyoto, Japan) with patients standing without shoes and heels against the wall. BP was measured on the right upper arm after 5 minutes of rest in a seated position using an electronic BP monitor (Omron HBP-9020; Omron Corporation, Kyoto, Japan). ICG data were collected by trained technicians at each visit in all patients, but ICG findings were not revealed in the standard arm to physicians or patients. ICG was performed with patients in the supine position, resting for 3 minutes before measurement. By applying a constant, low amplitude, high-frequency, alternating electrical current to the thorax, ICG device measures the corresponding voltage to detect beat-to-beat changes in thoracic electrical resistance, known as impedance, and with it stroke volume is estimated.^10, 11^ Then, using heart rate, mean arterial blood pressure, and BMI, other hemodynamic parameters are calculated, including CO, cardiac index (CI), SVR, systemic vascular resistance index (SVRI), arterial stiffness index (AS), and a volume parameter - thoracic blood saturation ratio (TBR).^12^ The ICG device used (CHM P2505, designed by Beijing Li-Heng Medical Technologies, Ltd, manufactured by Shandong Baolihao Medical Appliances, Ltd.) was developed based on improved hardware and advanced digital filtering algorithms,^13^ and has been validated versus both invasive thermodilution and non-invasive echocardiography in different settings.^14-16^

### Intervention

After randomization, therapy was initiated in all patients. Physicians in both groups were encouraged to prescribe medications consistent with published guidelines, their clinical judgement, and patient clinical characteristics. In the hemodynamic group, physicians were provided with patients’ ICG findings and a computerized clinical decision support of recommended treatment choices based on patients’ hemodynamic profiles. Specifically, the clinical decision support categorized patients into four clinically relevant hemodynamic phenotypes based on the value of CI, SVRI, heart rate, AS, and TBR.^17, 18^ These four hemodynamic phenotypes included cardiac phenotype (high HR or high CI), arterial vascular phenotype (high AS), peripheral vascular phenotype (high SVRI), and volemic phenotype (high TBR). Suggested treatment strategies were then provided for each phenotype (see details in **Figure 2**). Physicians were instructed to use this information to guide decisions about pharmacological agents and dosing. Physicians could share ICG information with patients in the hemodynamic arm. In the standard care group, physicians were not provided with patients’ ICG findings and were instructed to use their own clinical judgement to make treatment decisions. All patients in both groups received education on the importance of medication compliance

### Outcome measures

All patients were required to return to the clinic for a follow-up visit between 4 and 12 weeks after the baseline visit. During the follow-up visit, BP was measured on the right upper arm after 5 minutes of rest in a seated position using an electronic BP monitor (Omron HBP-9020; Omron Corporation, Kyoto, Japan). The technicians who measured BP were blinded to the intervention arm. The primary study end points were changes in SBP and DBP from baseline. Secondary study end points included (1) achievement of BP goal of <140/ 90 mmHg and (2) changes in SBP and DBP by baseline BP, age, sex, and BMI.

### Statistical analysis

We described continuous variables as mean ± SD and categorical variables as n (%). Differences in continuous variables between treatment groups were examined by the Student t test and in categorial variables using Fisher’s exact tests. Subgroup analysis was performed by baseline BP, age, sex, BMI, and hemodynamic phenotype. We used Breslow-Day test to test the consistency of different stratified odds ratio across subgroups and used Forest Plots for visualization. We performed additional evaluation of changes in hemodynamic parameters between baseline and follow-up visit by pair-sample t test. Statistical significance was defined as a 2-tailed P<0.05. All statistical analyses were conducted using R, version 3.4.1 (The R Foundation for Statistical Computing, Vienna, Austria).

## RESULTS

### Characteristics of study population

We screened 201 patients presenting to the hypertension clinic for outpatient care, from which we excluded 87 individuals whose baseline BP value is less than 140/90 mmHg, leaving 114 patients randomized to the intervention and control arms. We further excluded 12 patients who did not make follow-up visits within 4-12 weeks. Finally, a total of 102 patients (51 in the standard care group and 51 in the hemodynamic group) completed the study and were analyzed (**Figure 1**). Among 102 patients, the mean age of 54±14 years and 41% were female. Patients had a mean SBP of 150.9 (±11.5) mmHg, mean DBP of 91.1 (±11.3) mmHg, mean cardiac index of 3.1 (±0.7) L/min/m^2^, mean systemic vascular resistance index of 3017 (±731) dynes·sec/cm^5^/m^2^, mean heart rate of 72 (±10.6) beats/min (**Table 1**).

**Table 1.** Characteristics of study participants at baseline.

|  | Overall<br>(n=102) | Intervention group<br>(n=51) | Control group<br>(n=51) |
| --- | --- | --- | --- |
| Male, n (%) | 60 (59%) | 32 (63%) | 28 (55%) |
| Female, n (%) | 42 (41%) | 19 (37%) | 23 (45%) |
| Age, mean (SD) | 54 ± 14.0 | 55 ± 12.4 | 54 ± 15.5 |
| BMI (kg/m<sup>2</sup>) | 26.7 ± 3.8 | 26.4 ± 3.7 | 27.0 ± 3.9 |
| SBP (mmHg) | 150.9 ± 11.5 | 151.8 ± 12.6 | 150.0 ± 10.3 |
| DBP (mmHg) | 91.1 ± 11.3 | 92.7 ± 9.6 | 89.5 ± 12.6 |
| HR (BPM) | 72 ± 10.6 | 73 ± 11.2 | 70.0 ± 10.0 |
| CI (L/min/m<sup>2</sup>) | 3.1 ± 0.72 | 3.1 ± 0.64 | 3.0 ± 0.80 |
| AS (mmHg/mL/b) | 0.82 ± 0.33 | 0.82 ± 0.36 | 0.82 ± 0.30 |
| SVRI (dyn·s·m<sup>2</sup>/cm<sup>5</sup>) | 3017 ± 731 | 3057 ± 678 | 2975 ± 720 |
| TBR (%) | 0.78 ± 0.11 | 0.79 ± 0.11 | 0.76 ± 0.10 |
| Diabetes, n (%) | 25(25%) | 13(25%) | 12(24%) |
| CHD, n (%) | 4(4%) | 2(4%) | 2(4%) |
| Stoke, n (%) | 1(1%) | 1(2%) | 0(0%) |
| CKD, n (%) | 9(9%) | 7(14%) | 2(4%) |

The data of the two groups were not statistically significant ( $p>0.05$ for all). BMI: body mass index, SBP: systolic blood pressure, DBP, diastolic blood pressure, HR: heart rate, CI: cardiac output index, AS: aortic resistance index, SVRI: systemic vascular resistance index, TBR: thoracic blood volume saturation, CHD: coronary heart disease, CKD: chronic kidney disease.

**Figure 1.**
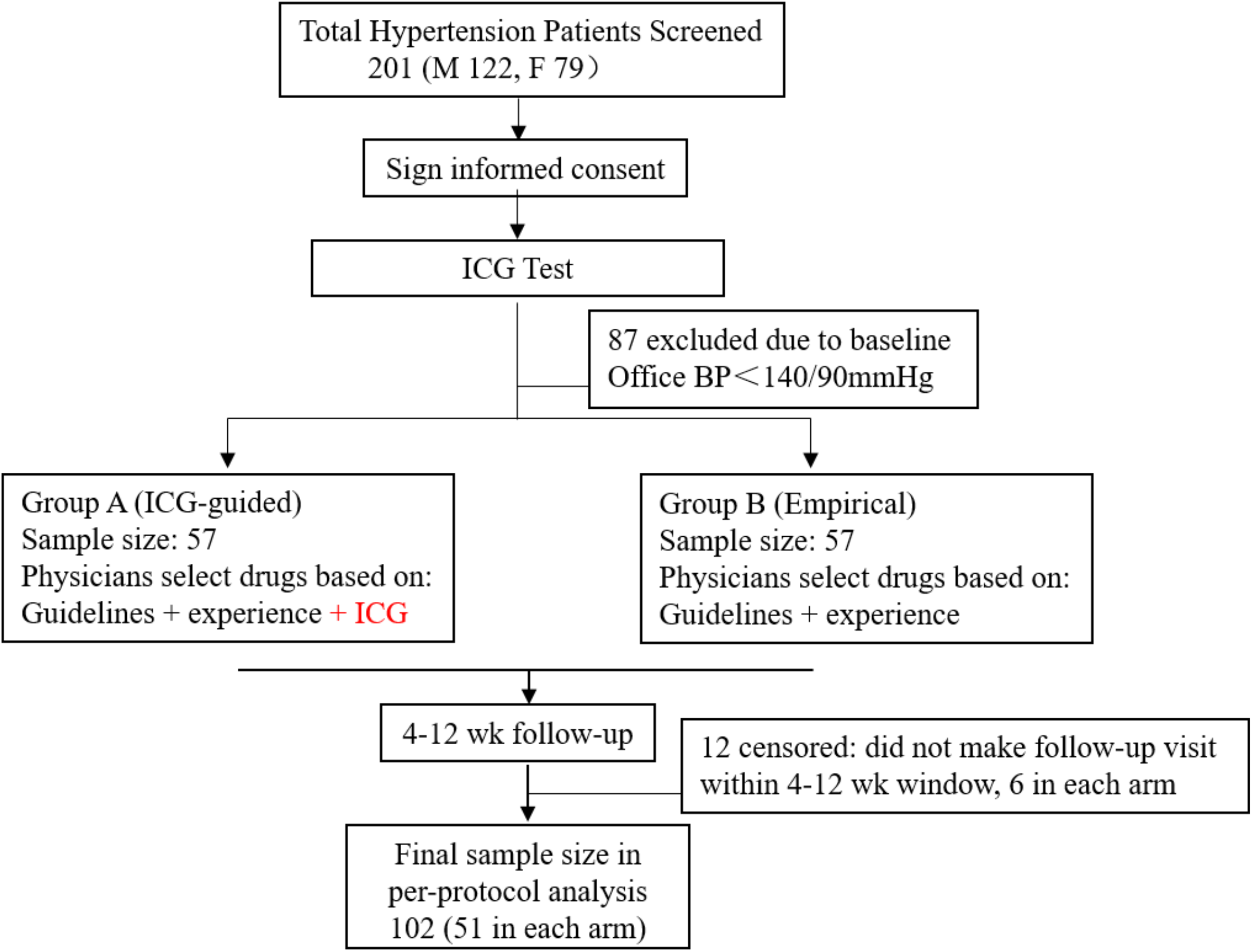
Enrolment and randomization of study participants.

**Figure 2.**
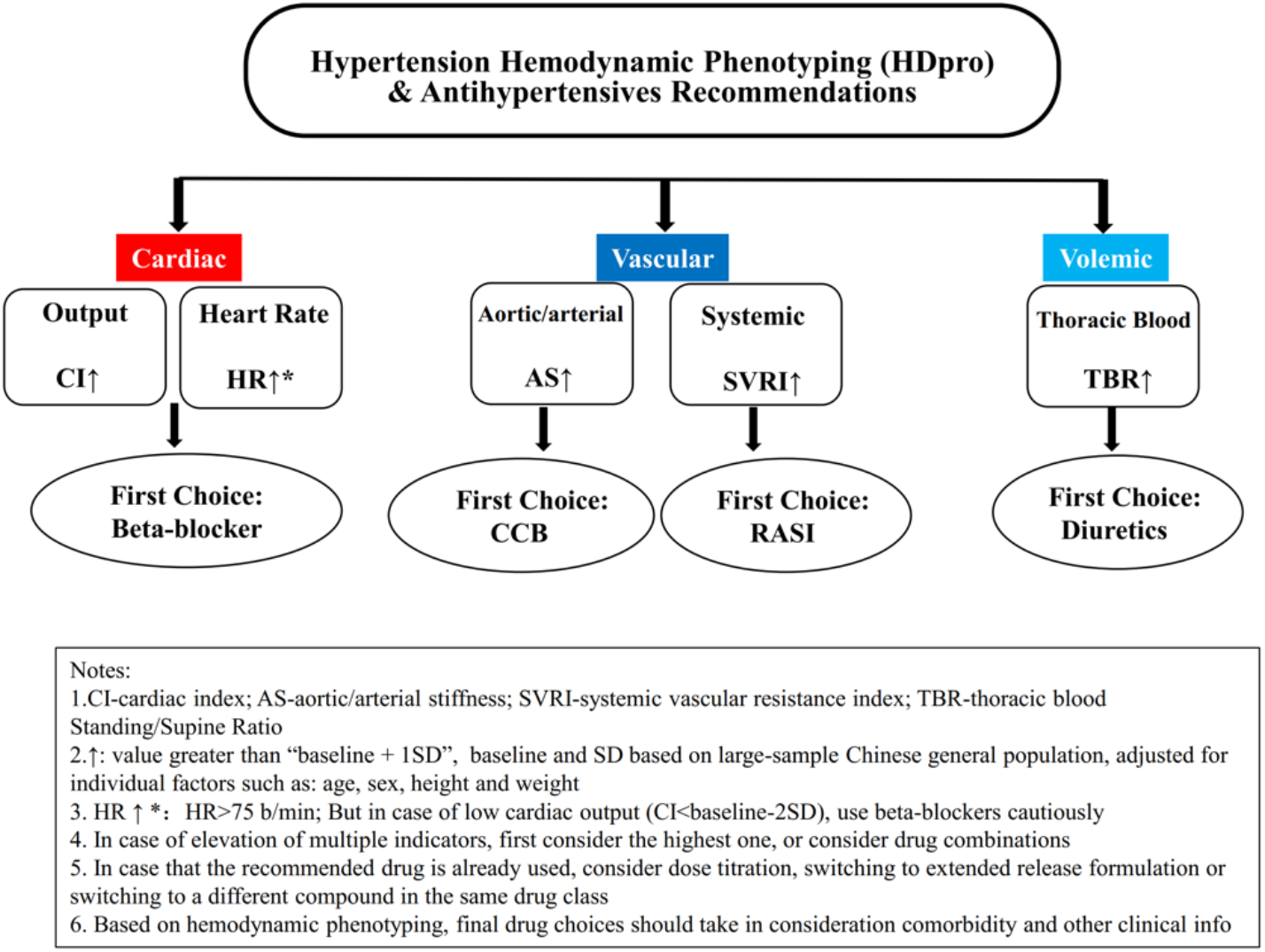
Suggested treatment strategy for the hemodynamic group.

In the hemodynamic group, 13 patients had cardiac phenotype (high HR or high CI), 11 had arterial vascular phenotype (high AS), 30 had peripheral vascular phenotype (high SVRI), and 17 volemic phenotype (high TBR), respectively. In the control group, 13 patients had cardiac phenotype, 18 had arterial vascular phenotype, 26 had peripheral vascular phenotype, and 11 volemic phenotype, respectively. There were no statistically significant differences in the number and class of antihypertensive medications, patient demographic, clinical, BP, or ICG variables at baseline between the hemodynamic group and the control group (**Table 1 and Table 2**).

**Table 2.** Number and class of medications prescribed at baseline and follow-up visit in hemodynamic and standard care groups.

|  | Baseline |  | *P*<br>*value* | Follow-up |  | *P*<br>*value* |
| --- | --- | --- | --- | --- | --- | --- |
|  | Intervention<br>group<br>(n=51) | Control<br>group<br>(n=51) |  | Intervention<br>group<br>(n=51) | Control<br>group<br>(n=51) |  |
| Antihypertensive medication (n±SE) | 1.39±0.18 | 1.41±0.16 | 0.988 | 2.37±0.16 | 2.41±0.18 | 0.812 |
| RASI | 41% | 45% | 0.842 | 76% | 84% | 0.455 |
| BB | 39% | 35% | 0.838 | 63% | 47% | 0.163 |
| CCB | 27% | 31% | 0.828 | 47% | 67% | 0.071 |
| DD | 31% | 29% | 1.000 | 52% | 35% | 0.161 |

RASI: renin-angiotensin system inhibitors, BB: beta-blockers, CCB: calcium channel blockers, DD: diuretics.

### Effect of the ICG-guided treatment strategy on blood pressure control

BP and ICG values at the baseline and follow-up visit as well as their differences between the two visits are shown in **Table 3 and Figure 3**. Both SBP and DBP reductions were significantly greater in the hemodynamic group from baseline to follow-up visit compared with the standard care group (SBP reductions: 19.9 ± 10.7 vs 12.0 ± 11.8 mmHg, P<0.001; DBP reduction: 11.3 ± 6.2 vs 4.9 ± 9.9 mmHg, P < 0.001). Final BP was lower in the hemodynamic group compared with the standard care group (SBP: 131.9 ±10.9 versus 138.0±13.7 mmHg, P< 0.001; DBP: 81.4 ±7.7 versus 84.6 ±12.9 mmHg, P< 0.001). The proportion of patients achieving BP goal of <140/ 90 mmHg was also larger in the hemodynamic group compared with the standard care group (67% versus 41%; P=0.017).

**Table 3.** BP reduction and achievement of BP goals in hemodynamic and standard care groups.

|  | N | Baseline BP<br>(mmHg) |  | Follow-up BP<br>(mmHg) |  | BP reduction<br>(mmHg) |  | Goal<br>achievement<br>rate (%) |
| --- | --- | --- | --- | --- | --- | --- | --- | --- |
|  |  | SBP | DBP | SBP | DBP | SBP | DBP |  |
| Intervention<br>group | 51 | 151.8 ±<br>12.6 | 92.7 ±<br>9.6 | 131.9 ±<br>10.9 | 81.4 ±<br>7.7 | -19.9 ±<br>10.7*** | -11.3 ±<br>6.2*** | 67** |
| Control<br>group | 51 | 150.0 ±<br>10.3 | 89.5 ±<br>12.6 | 138.0 ±<br>13.7 | 84.6 ±<br>12.9 | -12.0 ±<br>11.8 | -4.9 ±<br>9.9 | 41 |

SBP: systolic blood pressure, DBP: diastolic blood pressure, BP: blood pressure.
\*\* p<0.05, \*\*\*p<0.001

**Figure 3.**
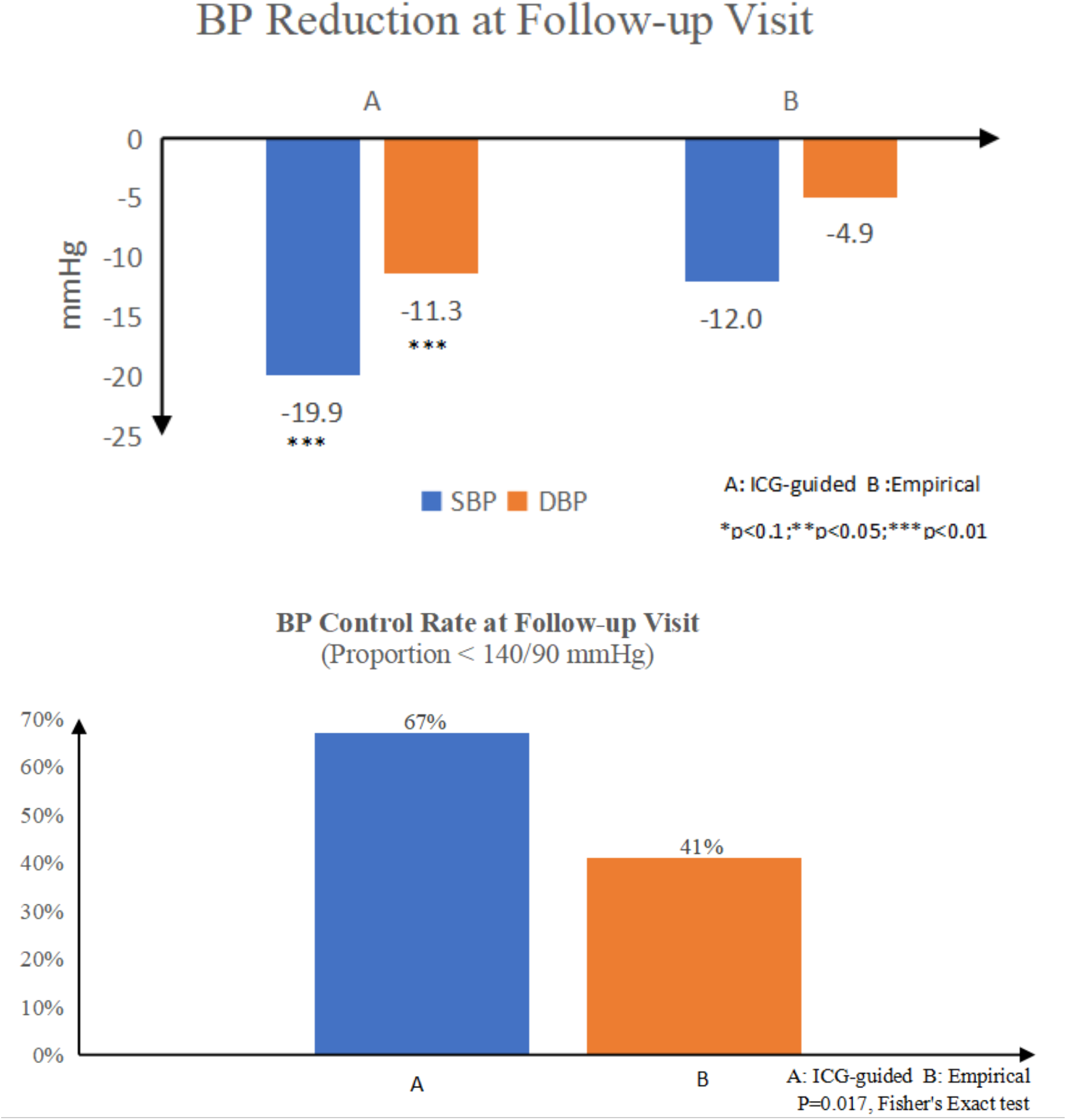
BP reduction and achievement of BP goals in hemodynamic and standard care groups.

Subgroup analyses by patient gender (men vs. women), age (≥50 years vs. <50 years), BMI (≥24 vs. <24 kg/m2), and baseline BP level (baseline SBP≥160 vs. 140-159 mmHg; baseline DBP≥90 vs. <90 mmHg) have consistently shown a greater BP reduction in the hemodynamic group compared with the standard care group. The differences between the two groups were statistically significant for all subgroups, except for DBP in men, SBP in age of <50 years, and DBP in BMI of <24 kg/m2 where the differences between the two groups were non-significant. The proportion of patients achieving BP goal of <140/ 90 mmHg was statistically significantly larger in the hemodynamic group compared with the standard care group for subgroups of men, age of <50 years, baseline SBP of <160 mmHg, and baseline DBP of ≥90 mmHg (**Figure 4**).

**Figure 4.**
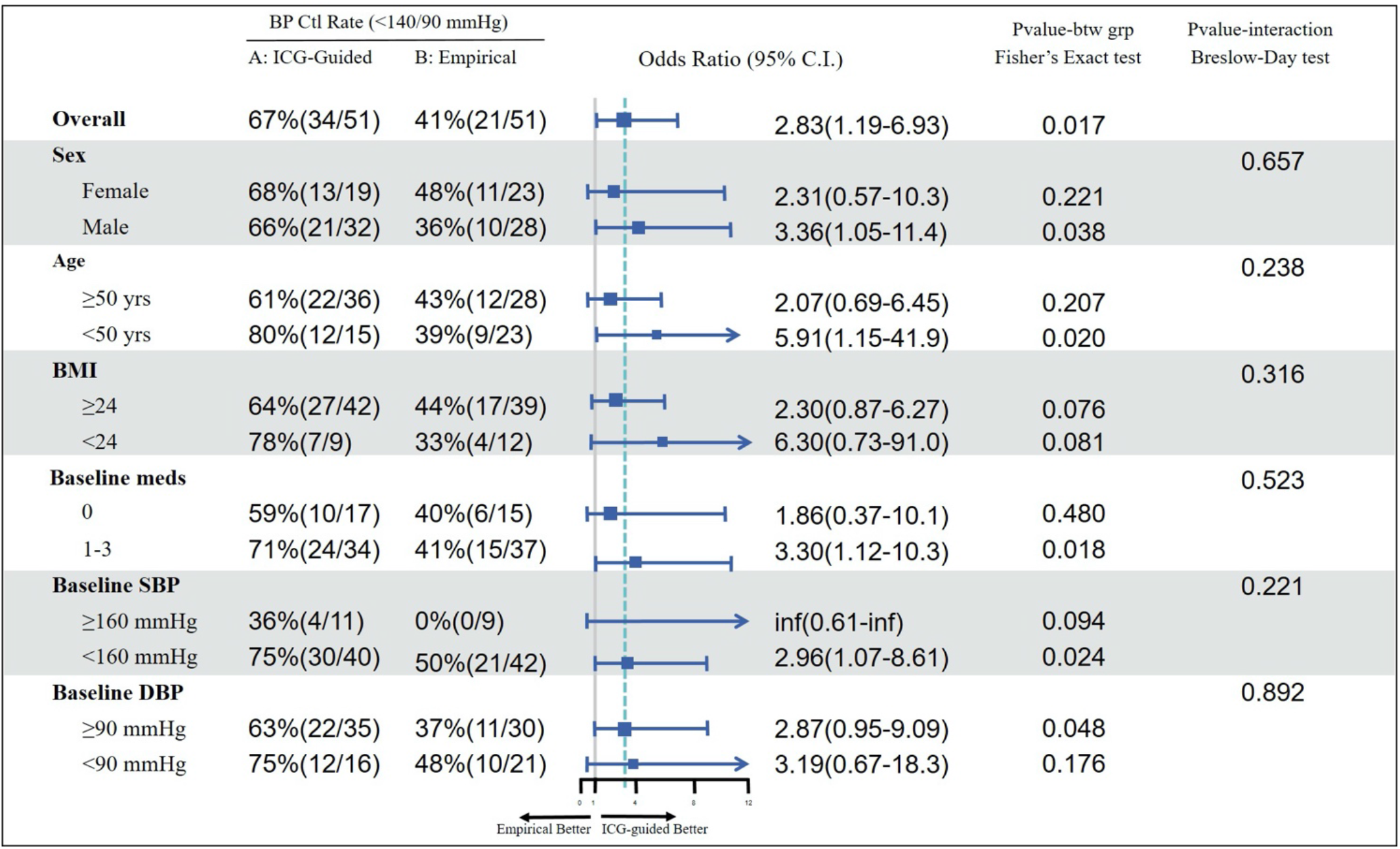
Achievement of BP goals by age, sex, BMI, baseline BP, use of medication at baseline.

Figure 5 showed BP reduction between two treatment groups by hemodynamic phenotypes. BP reduction was significantly larger in hemodynamic group compared with the standard care group for patients with hyperdynamic phenotype (high HR or high CI), arterial hyper-resistive phenotype (high AS), and peripheral artery hyper-resistive phenotype (high SVRI). BP reduction was not statistically significant in patients with high volume phenotype (high TBR).

**Figure 5.**
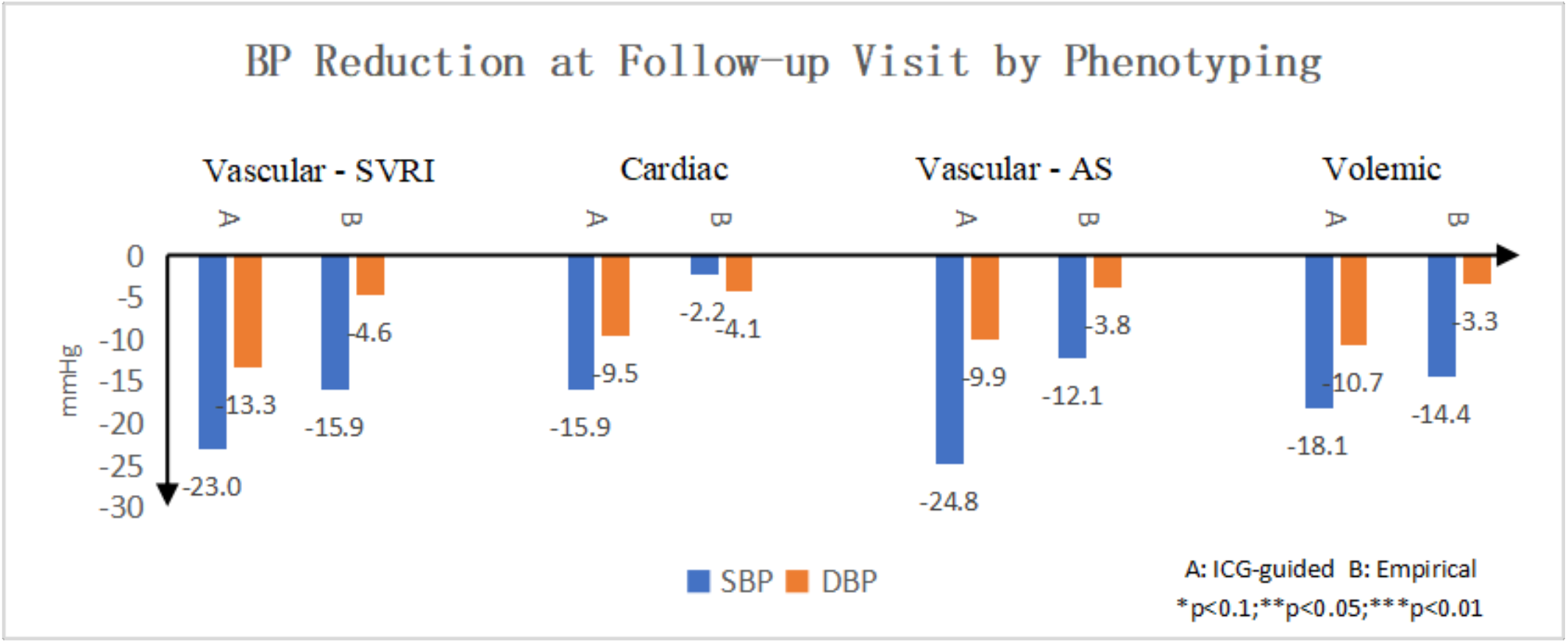
BP reduction by patients with different hemodynamic phenotypes.

### Correlation between antihypertensive agents and changes in hemodynamic parameters

In hemodynamic group, CI was statistically significantly reduced from baseline to follow-up visit in patients treated with beta-blockers (p = 0.044, **Figure 6**). TBR was statistically significantly reduced in patients treated with thiazide or thiazide-like diuretics (P =0.001). Both AS and SVRI were statistically significantly reduced in patients treated with calcium channel blockers (P =0.003), and SVRI was statistically significantly reduced in patients treated with renin–angiotensin system inhibitors (RASI, P < 0.001).

**Figure 6.**
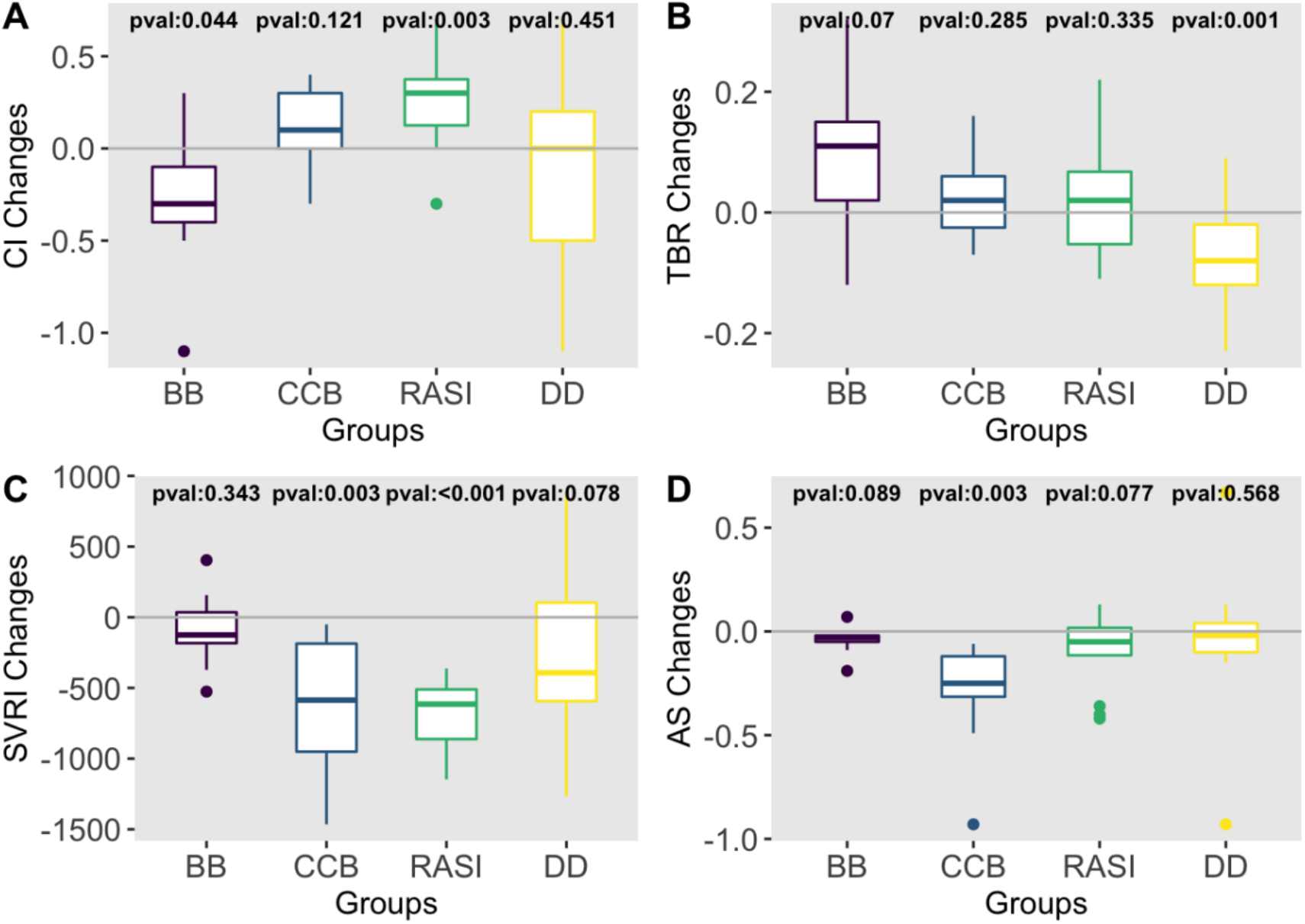
Impact of different antihypertensive agents on hemodynamic parameters.

## DISCUSSION

In a pilot trial of stable hypertensive patients routinely seen in clinical practice in China, we showed that an ICG-guided treatment strategy was more effective in reducing BP than standard therapy. These results were consistent across subgroups based on age, sex, BMI, baseline BP, and hemodynamic phenotype. Our findings suggest that antihypertensive therapy tailored to each patient’s hemodynamic abnormality could lead to more effective antihypertensive regimens and lay the groundwork for a more definitive trial.^19-22^

Of note, the reductions in BP in both groups were large, with an almost 20 mmHg decrease in SBP in the ICG-guided intervention group. The magnitude of the BP reduction in our study generally are consistent with previous studies conducted in the US. Smith et al conducted a randomized controlled trial of 164 uncontrolled hypertensive patients on 1 to 3 medications.^8^ After 3 months of treatment, patients in the ICG-guided group had an average SBP reduction of 19 mmHg compared with 12 mmHg in the standard care group. Taler et al randomized 104 patients with hypertension uncontrolled on two or more drugs to a 3-month trial of ICG-guided therapy or standard therapy directed by a hypertension specialist.^9^ In this study, the mean BP reduced from 169/87 mmHg to 139/72 mmHg in the ICG-guided group vs. from 173/91 mmHg to 147/79 mmHg in the control group. We further extended previous studies by using a pragmatic design to test the ICG-guided intervention in real-life clinical practice and conducting the study in a low- and middle-income country. We also provided a clinical decision support tool in addition to the ICG report to facilitate the antihypertensive treatment selection, producing a magnitude of BP improvement in routine clinical practice similar as that in the clinical trials.

There are several potential explanations for the findings in this study. First, the presumed mechanism for improved BP control with ICG-guided intervention is primarily due to personalized antihypertensive drug selection targeted at the hemodynamic cause of elevated BP. High BP results from one or more hemodynamic abnormality, including elevated CO, SVR, and blood volume.^23^ Different antihypertensive agents act on different mechanisms to reduce BP by reducing CO or SVR. For example, beta-blockers block the effects of the hormone adrenaline and make the heart to beat more slowly and with less force, which then reduce CO and lower BP. ACE inhibitors/ARBs interfere with the body’s renin-angiotensin-aldosterone system that leads to increased sodium and urine excreted, reduced resistance in blood vessels, and increased venous capacity, which then reduce SVR and lower BP. Our ICG-guided intervention provides data on the underlying cause of elevated BP and uses clinical decision support to guide clinicians in selecting antihypertensive therapies targeted at the hemodynamic abnormality associated with the elevated BP, thereby maximizing the BP lowering response for the given therapeutic selection.

Second, the larger reduction in BP in the intervention group may have been, in part, a reflection of improvement in therapeutic inertia. Therapeutic inertia, which refers to failure of clinicians to initiate or intensify treatments when the BP is not at goal, has been showed as a most common cause of uncontrolled BP in actively treated patients.^24, 25^ Providing clinician access to ICG findings of patients’ hemodynamic profiles and clinical decision support tool for treatment selection may reduce therapeutic inertia in the intervention group.

Finally, the improvement of BP may, at least in part, be associated with improved communications and shared decision making between the physician and the patient. The ICG report has served as a tool for physicians to communicate with and educate patients on the underlying hemodynamic abnormalities associated their high BP and rationale for antihypertensive therapy selection in the intervention group. Previous studies have reported that patient-physician communication is an integral part of clinical practice and patients who understand explanations from their physicians are more likely to acknowledge health problems, modify behavior, and adhere to medications accordingly.^26-28^

Our findings have important clinical implications. Current diagnosis and management of hypertension is primary based on degree of BP elevation alone, with little attention paid to the underlying hemodynamic profile. Our study provides evidence for better identification responders to a particular treatment regimen by profiling patients based on their hemodynamic profile using a simple, non-invasive test. As clinical care is moving towards precision medicine, our findings identify the needs of more refined hemodynamic measurement to facilitate personalized treatment in patients with hypertension. Additionally, the use of ICG-guided treatment strategy to achieve greater BP control offers a potential for better short-term use of healthcare resources. This is particularly relevant in low- and middle-income counties where resources to improve hypertension control are limited and need to be more efficiently used. Given hypertension affects over one billion adults (30% of the global adult population) in the world,^29^ such an approach has a large potential benefit in improving hypertension control and subsequently reducing a large number of cardiovascular events.

Several limitations should be considered in the interpretation of this study. First, this is a pilot study with limited number of participants and relatively short follow-up. We did not collect long-term follow-up data, which could have been useful to assess the long-term effect of ICG-guided treatment strategies in improving BP control. Second, our findings also warrant further study in other populations, as our study was conducted in a Chinese population and the results may not be generalizable to other populations. Third, we did not assess medication compliance among hypertensive patients, which may affect BP values of patients in the two arms. However, we used a pragmatic design to evaluate the effectiveness of interventions and we expect the medication compliance would be analogous to the scenarios in real-life routine clinical practice. Mediation refill rates were similar in the two arms as all patients fulfilled their prescriptions at the hospital pharmacy on the same day of the clinical encounters.

In conclusion, a treatment strategy guided by hemodynamic measurements reduced BP more effectively than standard care in this pilot trial in China. These findings justify further studies to provide more definitive evidence.

## Data Availability

Data are available upon request to the authors. Dr. Ningling Sun and Dr. Luyan Wang had access to all the study data.

## Reference

1. Oparil S, Zaman M and Calhoun DA. Pathogenesis of hypertension. Ann Intern Med. 2003;139:761–776.

2. Mahajan S, Gu J, Lu Y, Khera R, Spatz ES, Zhang M, Sun N, Zheng X, Zhao H, Lu H, Ma ZJ and Krumholz HM. Hemodynamic Phenotypes of Hypertension Based on Cardiac Output and Systemic Vascular Resistance. Am J Med. 2020;133:e127–e139.

3. Mahajan S, Gu J, Caraballo C, Lu Y, Spatz ES, Zhao H, Zhang M, Sun N, Zheng X, Lu H, Yuan H, Ma ZJ and Krumholz HM. Relationship of Age With the Hemodynamic Parameters in Individuals With Elevated Blood Pressure. Am Geriatr Soc. 2020.

4. Drazner MH, Thompson B, Rosenberg PB, Kaiser PA, Boehrer JD, Baldwin BJ, Dries DL and Yancy CW. Comparison of impedance cardiography with invasive hemodynamic measurements in patients with heart failure secondary to ischemic or nonischemic cardiomyopathy. Am J Cardiol. 2002;89:993–5.

5. Sageman WS, Riffenburgh RH and Spiess BD. Equivalence of bioimpedance and thermodilution in measuring cardiac index after cardiac surgery. J Cardiothorac Vasc Anesth. 2002;16:8–14.

6. Ferrario CM and Smith RD. Cost-effectiveness of impedance cardiography testing in uncontrolled hypertension. Am Heart Hosp J. 2006;4:279–289.

7. Ferrario CM, Flack JM, Strobeck JE, Smits G and Peters C. Individualizing hypertension treatment with impedance cardiography: a meta-analysis of published trials. Ther Adv Cardiovasc Dis. 2010;4:5–16.

8. Smith RD, Levy P, Ferrario CM and Consideration of Noninvasive Hemodynamic Monitoring to Target Reduction of Blood Pressure Levels Study G. Value of noninvasive hemodynamics to achieve blood pressure control in hypertensive subjects. Hypertension. 2006;47:771–7.

9. Taler SJ, Textor SC and Augustine JE. Resistant hypertension: comparing hemodynamic management to specialist care. Hypertension. 2002;39:982–8.

10. Bernstein DP. A new stroke volume equation for thoracic electrical bioimpedance: theory and rationale. Crit Care Med. 1986;14:904–9.

11. Sherwood A, Allen MT, Fahrenberg J, Kelsey RM, Lovallo WR and van Doornen LJ. Methodological guidelines for impedance cardiography. Psychophysiology. 1990;27:1–23.

12. Yancy C and Abraham WT. Noninvasive hemodynamic monitoring in heart failure: utilization of impedance cardiography. Congest Heart Fail. 2003;9:241–50.

13. Ma L. Development and application of the latest model of the cardiac hemodynamics monitoring system. International J Cardiovasc Med. 2009;10:11.

14. An X-g, Zhao Y and Gao J. Clinic evaluation of noninvasive hemodynamic monitoring in patients undergoing coronary artery surgery. Chin J Cardiovasc Rev. 2008;2:010.

15. Hong H, Jin X, Pan C, Gao X, Liu M, Jiang H, Ge J. Analysis of the correlation between non-invasive hemodynamic monitor and cardiac echocardiography on the evaluation of cardiac function. Chinese Journal of Medical Instrumentation. 2009;33:328–331.

16. Chen W, Huang D, Zeng J, Deng R, Deng G, Chen W, Zou Y. Application of noninvasive cardiac hemodynamic monitor for ICU critically ill patients. Hainan Medical Journal. 2018;29:15.

17. Wannenburg T and Little WC. Regulation of cardiac output. In: A. Jeremias and D. L. Brown, eds. Cardiac Intensive Care Second ed. Philadelphia: W.B. Saunders; 2010: 61–68.

18. Gidwani UK, Mohanty B and Chatterjee K. The pulmonary artery catheter: a critical reappraisal. Cardiol Clin. 2013;31:545-65, viii.

19. Smith RD, Levy P and Ferrario CM. Value of noninvasive hemodynamics to achieve blood pressure control in hypertensive subjects. Hypertension. 2006;47:771–7.

20. Ventura HO, Taler SJ and Strobeck JE. Hypertension as a hemodynamic disease: The role of impedance cardiography in diagnostic, prognostic, and therapeutic decision making. Am J Hypertens. 2005;18:26S–43S.

21. Taler SJ, Textor SC and Augustine JE. Resistant hypertension: Comparing hemodynamic management to specialist care. Hypertension. 2002;39:982–988.

22. Sanford T, Treister N and Peters C. Use of noninvasive hemodynamics in hypertension management. Am J Hypertens. 2005;18:87S–91S.

23. Mayet J and Hughes A. Cardiac and vascular pathophysiology in hypertension. Heart. 2003;89:1104–1109.

24. O’Connor PJ. Overcome clinical inertia to control systolic blood pressure. Arch Intern Med. 2003;163:2677–8.

25. Okonofua EC, Simpson KN, Jesri A, Rehman SU, Durkalski VL and Egan BM. Therapeutic inertia is an impediment to achieving the Healthy People 2010 blood pressure control goals. Hypertension. 2006;47:345–51.

26. Travaline JM, Ruchinskas R and D’Alonzo GE, Jr. Patient-physician communication: why and how. J Am Osteopath Assoc. 2005;105:13–8.

27. Bull SA, Hu XH, Hunkeler EM, Lee JY, Ming EE, Markson LE and Fireman B. Discontinuation of use and switching of antidepressants: influence of patient-physician communication. Jama. 2002;288:1403–9.

28. Ciechanowski PS, Katon WJ, Russo JE and Walker EA. The patient-provider relationship: attachment theory and adherence to treatment in diabetes. Am J Psychiatry. 2001;158:29–35.

29. Collaboration NCDRF. Worldwide trends in blood pressure from 1975 to 2015: a pooled analysis of 1479 population-based measurement studies with 19.1 million participants. Lancet. 2017;389:37–55.

